# Nutrition Policies in Germany: A Systematic Assessment with the Food Environment Policy Index (Food-EPI)

**DOI:** 10.1101/2021.10.11.21264774

**Authors:** Peter von Philipsborn, Karin Geffert, Carmen Klinger, Antje Hebestreit, Jan Stratil, Eva Rehfuess, for the PEN Consortium

## Abstract

**Objective:** To systematically assess Germany’s nutrition policies, to benchmark them against international best practices, and to identify priority policy actions to improve population-level nutrition in Germany.

**Design:** We applied the Food Environment Policy Index (Food-EPI), a methodological framework developed by the INFORMAS network. Qualitative content analysis of laws, directives and other documents formed the basis of a multi-staged, structured consultation process.

**Setting:** Germany.

**Participants:** The expert consultation process included 55 experts from academia, public administration, and civil society.

**Results:** Germany lags behind international best practices in several key policy areas. For 18 policy indicators, the degree of implementation compared to international best practices was rated as very low, for 21 as low, for 8 as intermediate, and for none as high. In particular, indicators on food taxation, regulation of food marketing, and retail and food service sector policies were rated as very low to low. Identified priority actions included the binding implementation of nutrition standards for schools and kindergartens, a reform of the value added tax on foods and beverages, a sugar-sweetened beverage tax and stricter regulation of food marketing directed at children.

**Conclusions:** The results show that Germany makes insufficient use of the potential of evidence-informed health-promoting nutrition policies. Adopting international best practices in key policy areas could help to reduce the burden of nutrition-related chronic disease and related inequalities in nutrition and health in Germany. Implementation of relevant policies requires political leadership, a broad societal dialogue, and evidence-informed advocacy by civil society, including the scientific community.

## Introduction

Unhealthy dietary patterns are among the most important preventable risk factors for disease and premature death worldwide (1). In particular, unhealthy diets can increase the risk for obesity, type 2 diabetes mellitus, cardiovascular disease, and certain cancers (1, 2). Approximately 8% of the global burden of disease is attributed to dietary risks (1, 3). Over the past decades, global dietary patterns have undergone fundamental shifts, with an increase in the per-capita consumption of ultra-processed foods and beverages with a high content of sugar, refined grains, fat, and salt (4-6). These shifts have contributed to a rising burden of diet-related chronic disease (7). In particular, the global prevalence of obesity has doubled from 6% to 13% among adults between 1985 and 2016 (8), and risen seven-fold from 1% to 7% among children between 1975 and 2016 (9). Similarly, the global prevalence of type 2 diabetes mellitus has roughly doubled from 5% to 9% between 1980 and 2014 (10).

Individual dietary intake is strongly influenced by the food environment, defined as “the collective physical, economic, policy and sociocultural surroundings, opportunities and conditions that influence people’s food and beverage choices and nutritional status” (11). The food environment is shaped, among others, by public policies in areas such as food composition, labelling, taxation, marketing, and public procurement. The evidence on the effectiveness of public policies supporting healthy diets has grown considerably over the past years but implementation remains highly uneven across countries (12-14).

Unhealthy diets and diet-related diseases are also a key public health issue in Germany. In Germany’s National Nutrition Survey II (NVS II), conducted in a nationally representative sample from 2005 to 2007, for most food groups significant deviations from the recommended consumption were found (15). Follow-up studies conducted from 2008 to 2015 as part of the longitudinal National Nutrition Monitoring (NEMONIT) did not detect relevant improvements over time (16). Consumption levels of vegetables, meat and sausages, and dairy products remained largely unchanged, while fruit consumption slightly decreased and confectionery consumption increased (16). The prevalence of obesity in Germany is 23% among adults and 6% among children, and is therefore above the European average (17). While the prevalence of obesity has remained stable among children since 2003 (18), it continues to increase among adults, in particular among low socio-economic status groups (19). Obesity and its underlying dietary risk factors are also associated with high costs for the health care system. For instance, direct health-related costs of excessive consumption of sugar, fat, and salt in Germany were estimated to account for nearly 17 billion euros, or 7% of all direct health care costs in 2008 (20).

Against this background, the German government has implemented or announced a number of measures to support healthier diets on the population level (21, 22). These include a National Reformulation and Innovation Strategy for Sugar, Salt, and Fat in Processed Foods; the introduction of the Nutri-Score nutrition labelling system on a voluntary basis; the founding of a new national nutrition education and information centre (Bundeszentrum für Ernährung, or BZfE); and measures to improve the quality of food served in schools and kindergartens through information and training (21-23).

To the authors’ knowledge, the current nutrition-related policy landscape in Germany has, however, not yet been assessed in a comprehensive, systematic and international comparative manner. Internationally, a number of approaches for the comprehensive assessment of the degree of implementation of nutrition policies have been proposed (11, 24, 25). One of most widely used approaches is the Food Environment Policy Index (Food-EPI), which has been developed by the international scientific network INFORMAS (International Network for Food and Obesity/non-communicable Diseases Research, Monitoring and Action Support) based on the review and evaluation of existing approaches and the involvement of stakeholders from policy and practice (11, 26, 27). To date, the Food-EPI has been applied in around 40 countries worldwide (28). In the present paper, we use the Food-EPI framework systematically assess the current nutrition policy landscape in Germany, benchmark it against international best practices for health-promoting nutrition policies, and identify priority actions for reform. This work was conducted within the Policy Evaluation Network (PEN, www.jpi-pen.eu) as a project of the Joint Programming Initiative “A Healthy Diet for a Healthy Life” (29).

## Methods

The Food-EPI framework and its development have been described in greater detail elsewhere (11). In short, the Food-EPI is a policy analysis framework based on a qualitative content analysis of relevant documents, as well as a structured, multi-stage expert consultation process. It includes 13 domains, 7 of which describe substantive policy areas (such as labelling and taxation), while 6 describe overarching structures and supportive functions (such as monitoring and surveillance, governance, and funding). For each domain, 2 to 5 indicators are defined, resulting in a total of 47 indicators (see table 1 for an overview, and the supplementary appendix for a more detailed description). The focus is on policies relevant for the promotion of healthy diets that minimize the risk of chronic, nutrition-related conditions such as obesity and type 2 diabetes mellitus. Aspects of food safety (i.e. prevention of microbiological, chemical or physical contaminations and food-borne infections) are excluded, as are the promotion of breast feeding and the regulation of dietary supplements. Aspects of environmental sustainability and animal welfare are also not covered.

**Table 1:**
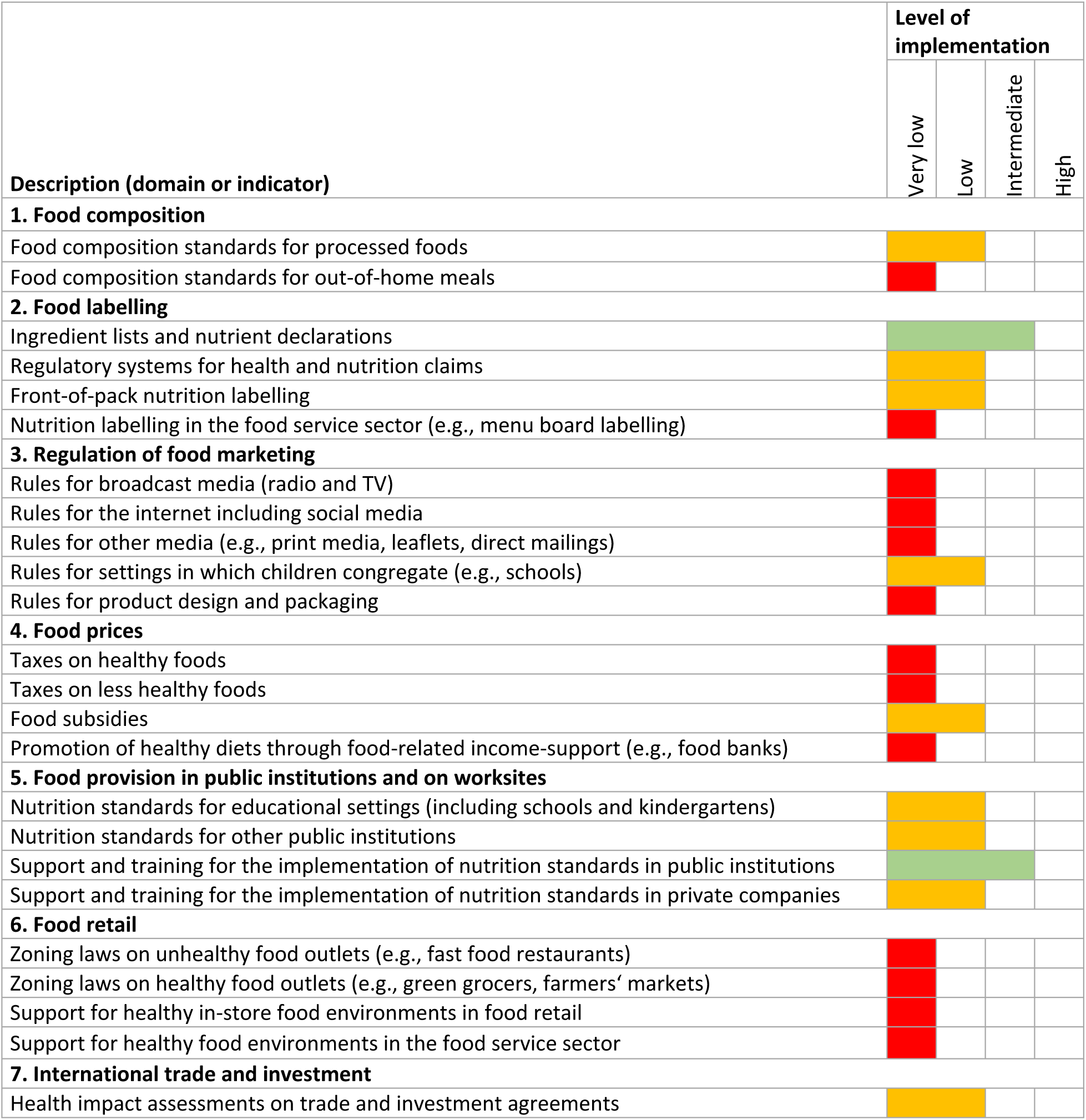
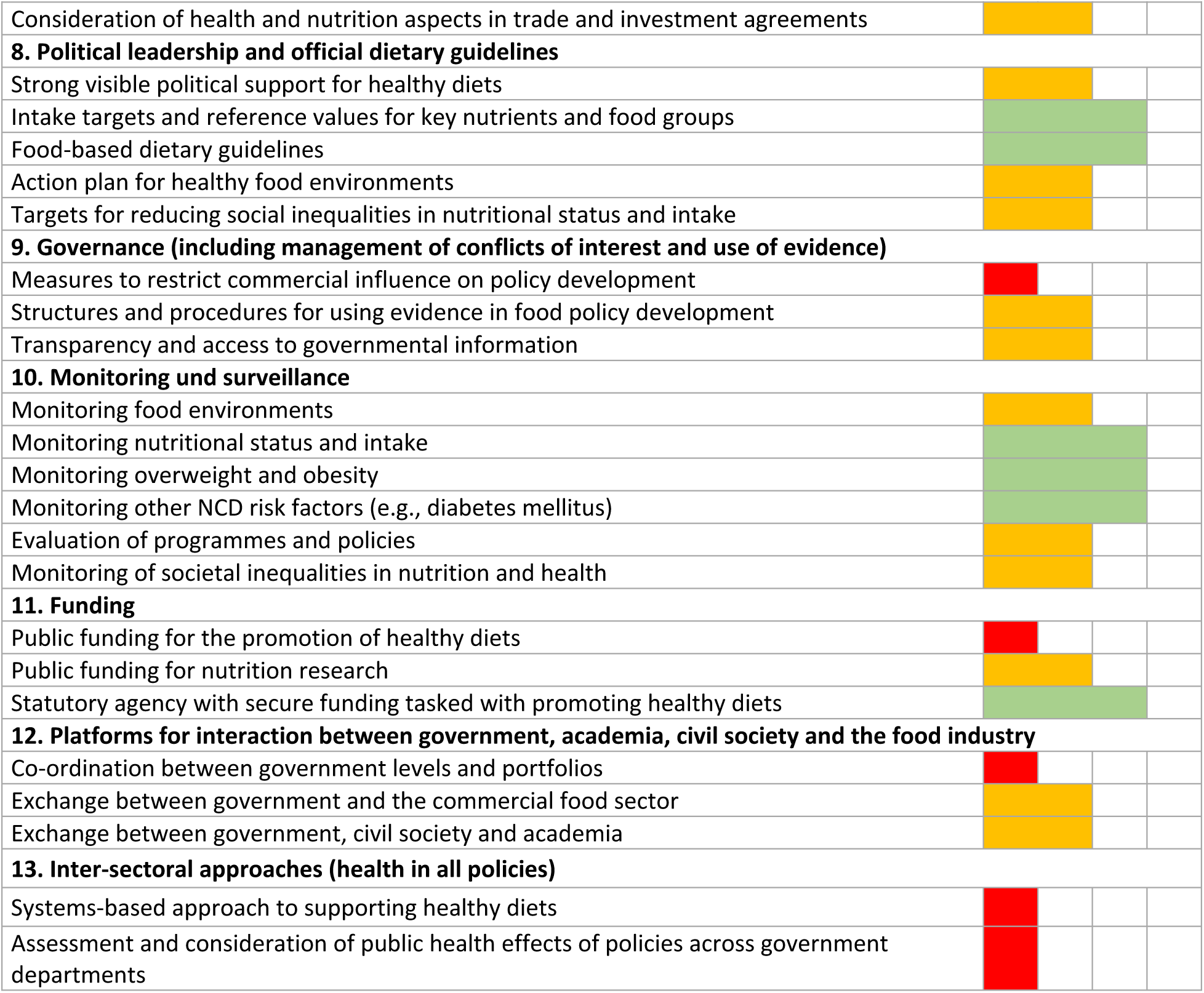
Benchmarking results for Germany. Results show the experts’ assessment of the degree of implementation in Germany relative to international best practices. A detailed description of the current implementation in Germany and international best practices was published separately (31).

The country-level implementation of the Food-EPI involves the following steps, described in greater detail in the subsequent paragraphs:

1. Adaptation of the international Food-EPI framework to the national context
2. Identification of relevant policies and compilation of descriptive information on these in an evidence report
3. Establishment of an expert panel, including researchers and representatives of relevant government bodies and civil society organisations
4. Validation of the evidence report by the expert panel
5. Assessment of the degree of implementation for each indicator in comparison with international best practice examples by the expert panel (benchmarking)
6. Identification and prioritization of policy recommendations by the expert panel

In conducting and reporting our research, we followed the Consolidated Criteria for Reporting Qualitative Studies (COREQ) (30). The COREQ Checklist is provided in table s5 in the supplementary appendix. Ethics approval was granted by the ethics committee of the LMU Munich.

### Adaptation of the Food-EPI to the national context

We identified 6 indicators in the international Food-EPI framework that we found to be largely overlapping in the German context, and which we therefore merged into three pairs, as shown in table s4 in the supplementary appendix.

### Identification of relevant policies and compilation of descriptive information

To identify relevant policies, we searched for policy-related, publicly available documents (such as laws, directives, official reports and other government documents, position statements, press releases, and minutes of parliamentary debates). Any document providing substantive information on any of the Food-EPI indicators was included. We searched the following websites manually, with Google Advanced Search and through site-specific search functions, using search terms related to nutrition and obesity prevention:

- The websites of federal ministries, regulatory agencies, and administrative bodies in the areas of nutrition and health (i.e. the Federal Ministry of Health, the Federal Ministry of Nutrition and Agriculture, the Federal Centre for Nutrition, the Robert Koch Institute, the Max Rubner Institute and the Federal Centre for Risk Assessment)
- The website of the Bundestag, Germany’s parliament, and its archive of parliamentary debates, motions and inquiries
- The websites of non-governmental organizations, including professional organizations, trade associations and pressure groups
- German and European legal databases (N-Lex and EU-LEX)
- International policy databases maintained by the World Health Organization (WHO) (i.e. the WHO NCD Document Repository, the Global Database on the Implementation of Nutrition Action, and the e-Library of Evidence for Nutrition Actions) and scientific associations (the NOURISHING database by the World Cancer Research Fund International, and Global Obesity Observatory by World Obesity)

We used the data analysis software MAXQDA to conduct a deductive qualitative content analysis of the included documents, using the domains and indicators of the Food-EPI as coding framework. Data was coded independently by two authors ([author initials hidden for peer review]). Results were then summarized narratively, and, where appropriate, illustrated with quotes. The information gathered through this analysis was complemented by consulting existing literature (e.g., textbooks on German and European food law, and academic publications on nutrition policies in Germany), and through targeted Google searches. All information was fully referenced to ensure transparency. The process was guided by the aim to provide a comprehensive, yet succinct description of the current legal framework as well as existing government policies, programmes, projects and initiatives for each of the 47 Food-EPI indicators.

In the process of data collection and analysis, we focused on the federal (i.e. national) level in Germany. However, we also included information on EU regulations and programmes applicable to Germany – complemented, where available, by information on how these are implemented nationally. In policy areas where the main regulatory authority lies with the subnational (state or local) level, we aimed to provide an overview of relevant policies on these levels, too, e.g., by stating how many of Germany’s 16 states have policies on a specific issue, or by providing illustrative examples.

The information gathered was compiled in an evidence report, including references to all sources used (31).

### Recruitment of an expert panel

For the subsequent steps, we recruited an expert panel, including academics and experts from professional and scientific associations as well as civil society and governmental organizations. To avoid conflicts of interest, no industry representatives were involved. We used purposive sampling to identify relevant experts, based on the authors’ knowledge, research and contacts, as well as recommendations by included experts. Experts were contacted by email and provided with an information sheet (see supplementary appendix).

### Validation of the evidence report

To ensure that the information compiled in the evidence report is complete, accurate and up to date, we sent a draft to the members of the expert panel, asking them to provide feedback in the form most convenient to them (e.g., using the comment and track-changes mode of their word processor, or by email or telephone). We then revised the draft evidence report based on their feedback.

### Benchmarking

The next step involved the benchmarking, i.e. the assessment, for each indicator, of the degree of implementation in Germany as compared to international best practices. The selection of international best practice examples was based on a consensual collection produced by INFORMAS and updated within the Policy Evaluation Network. For application in Germany, a selection of relevant examples was taken from this collection by the authors and complemented with examples from the NOURISHING database of the World Cancer Research Fund International (32).

For the benchmarking, the participating experts were provided with a table allowing, for each of the 47 indicators, a direct comparison of existing policies in Germany (as identified and described in the evidence report) with the international best practice examples. The experts could rate the degree of implementation in Germany on a four-point Likert scale as very low, low, medium, or high. The rating could be completed on paper or online, with the online version allowing for anonymous data submission. For a summary assessment of the degree of implementation, we calculated the median of the ratings provided by the participating experts for each indicator.

### Identification and prioritization of policy recommendations

We searched existing catalogues of recommendations by the WHO and national professional and scientific associations, and compiled a draft list of relevant policy recommendations for Germany. This list was sent to the members of the expert panel for feedback and suggestions on additional policy recommendations. Subsequently, a revised list of recommendations was discussed with the expert panel during a 2-hour online workshop. We recorded and transcribed the workshop, using the transcript for further revisions of the draft list of recommendations. Analogous to the structure of the Food-EPI framework, recommendations were divided into two areas: policy actions, such as a tax on sugar-sweetened beverages (SSB), and infrastructure support actions, such as improved monitoring and surveillance.

Based on the final list of recommendations, the expert panel prioritized policy actions and infrastructure support actions according to two criteria – i) their contribution to improving nutrition at the population level, and ii) their achievability, defined as the likelihood of policy adoption and the feasibility of legal and administrative implementation in Germany. Policy actions were additionally prioritized according to a third criterion – iii) their contribution to improving nutrition in socially disadvantaged population groups. For the prioritization, each recommendation could be given up to 5 points per criterion, with the total number of points to be distributed per criterion limited to twice the number of recommendations. For example, there were 10 recommendations on infrastructure support actions, implying that participating experts could distribute up to 20 points across the 10 recommendations for the criterion of impact, and 20 points for the criterion of achievability. For data analysis, we calculated the arithmetic mean of the points awarded by the participating experts to each recommendation for each criterion. To arrive at an overall ranking of recommendations, we calculated a summary score for each recommendation by summing up the scores for the individual criteria.

## Results

### Expert response rate and composition of expert panel

We contacted 72 experts, 55 of whom agreed to participate. A list of participating experts including their affiliations is provided in table s1 of the supplementary appendix. 53% (n=29) of participating experts were based in academia, 27% (n=15) in professional and scientific associations and civil society organizations, and 20% (n=11) in government agencies. One participant reported receiving lecture and reviewer fees from food companies as a relevant conflict of interest; for the others, no relevant conflicts of interest were identified. Of the 55 participating experts, 91% (n=50) provided feedback on the evidence report, 67% (n=37) participated in the benchmarking of policies, and 53% (n=29) in the prioritization of policies.

### Existing policies and their benchmarking against international best practices

A detailed description of existing policies in Germany based on a total of 341 documents reviewed and analysed, as well as the international best practice examples used in the benchmarking exercise, were published separately in German (31). Table 1 presents results of the benchmarking across the 13 Food-EPI domains, i.e. food composition, food labelling, regulation of food marketing, food prices, food provision in public institutions and on worksites, food retail, international trade and investment, political leadership and official dietary guidelines, governance, monitoring and surveillance, funding, platforms for interaction between government, academia, civil society and the food industry and inter-sectoral approaches. For 18 indicators, the degree of implementation in Germany relative to international best practices was rated as very low, for 21 as low, for 8 as intermediate, and for none as high. The lowest ratings were observed for regulation of food advertisement and marketing, food pricing, promotion of healthy food choices in retail settings, and intersectoral approaches. The highest ratings were noted for political leadership and official dietary guidelines and monitoring and surveillance.

### Identification and prioritization of policy recommendations

We identified a total of 18 recommendations on policy actions, and 10 recommendations on infrastructure support actions (see tables 2 and 3). A detailed description of these is provided in tables s2 and s3 in the supplementary appendix, including a full list of references. The following policy actions were ranked highest with regard to their impact on improving population-level nutrition (see table 2): a health-promoting reform of the value added tax (VAT) (i.e. lower tax rates on healthier foods and beverages, and higher rates on less healthy ones); mandatory nutrition standards for schools and kindergartens; regulation of marketing of unhealthy foods and beverages towards children; and a tax on SSB. Regarding achievability, the following policy measures were ranked highest: mandatory nutrition standards for schools and kindergartens; an action plan on the promotion of drinking water; and nutrition education in schools and kindergartens (see table 2). Figure 1 illustrates that some of the policy actions with the greatest potential for improving population-level nutrition were assessed to be relatively more achievable (e.g. mandatory nutrition standards for schools and kindergartens), whereas the implementation of others is likely to be more challenging (e.g., a health promoting VAT, and mandatory reformulation of processed foods). Most policies with a potential to improving population-level nutrition were also considered to be likely to reduce inequalities in nutrition and health.

**Table 2:**
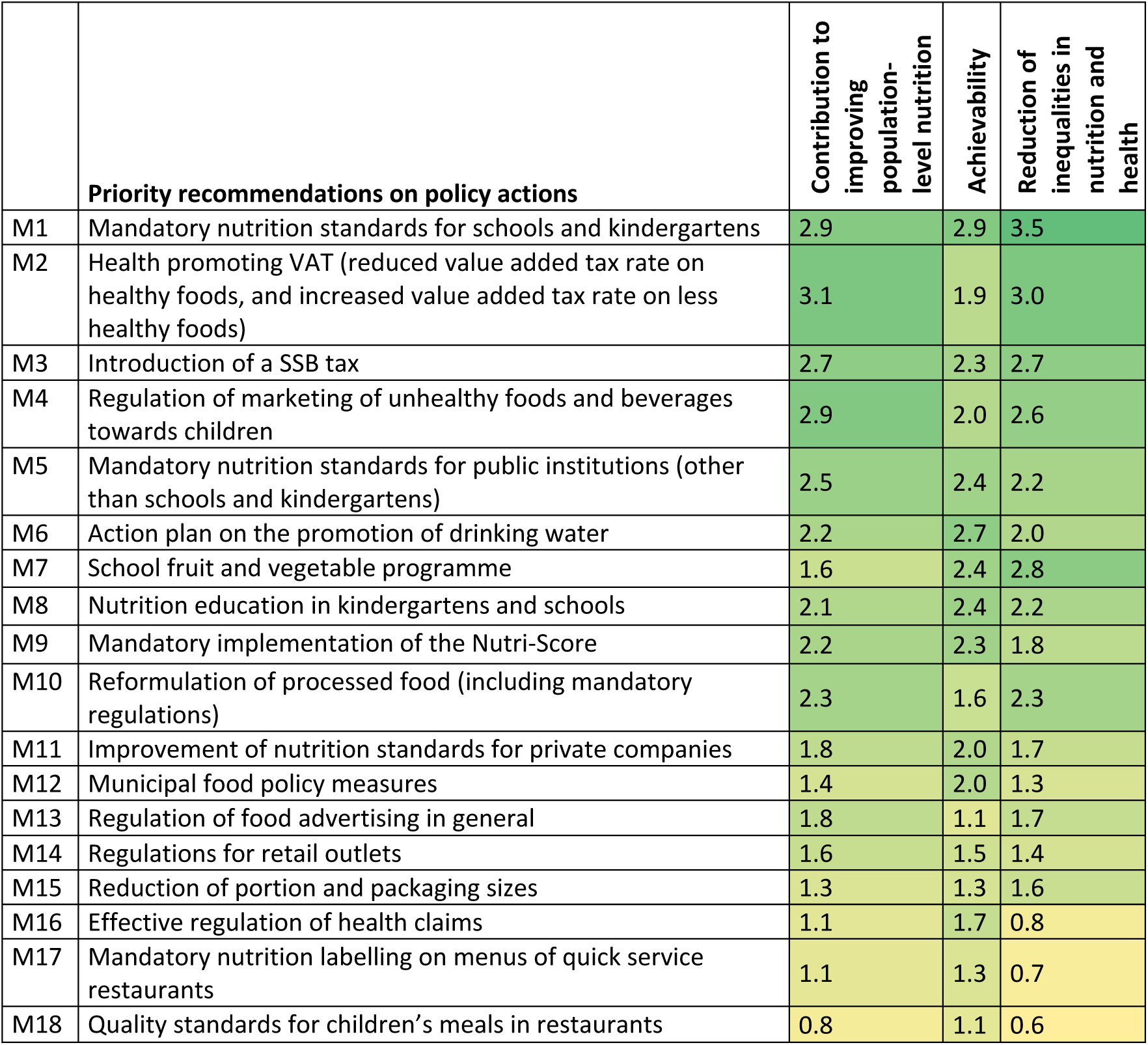
Priority recommendations on policy actions to improve population-level nutrition in Germany, based on expert judgement. Higher scores/darker colours indicate a higher priority.

**Table 3:** Priority recommendations for infrastructure support actions to improve population-level nutrition in Germany, based on expert judgement. Higher scores indicate a higher priority.

|  |  | Contribution to improving population-level nutrition | Achievability |
| --- | --- | --- | --- |
| **Priority recommendations for infrastructure support actions** |  |  |  |
| S1 | Evaluation of interventions and policies | 2.7 | 2.3 |
| S2 | Monitoring of nutritional status and dietary behaviour | 2.1 | 2.0 |
| S3 | Knowledge transfer between policy, practice and research | 2.1 | 1.9 |
| S4 | Nutrition education in the curricula of relevant professions | 1.9 | 2.0 |
| S5 | Monitoring of food environments | 2.0 | 1.9 |
| S6 | Local networks and initiatives | 1.6 | 2.2 |
| S7 | Action plan for healthy nutrition | 1.6 | 2.0 |
| S8 | Nutrition policy capacity building | 1.9 | 1.6 |
| S9 | Improving the availability of data | 1.6 | 1.7 |
| S10 | Capacities for the further development of nutrition recommendations | 1.6 | 1.7 |

**Figure 1:**
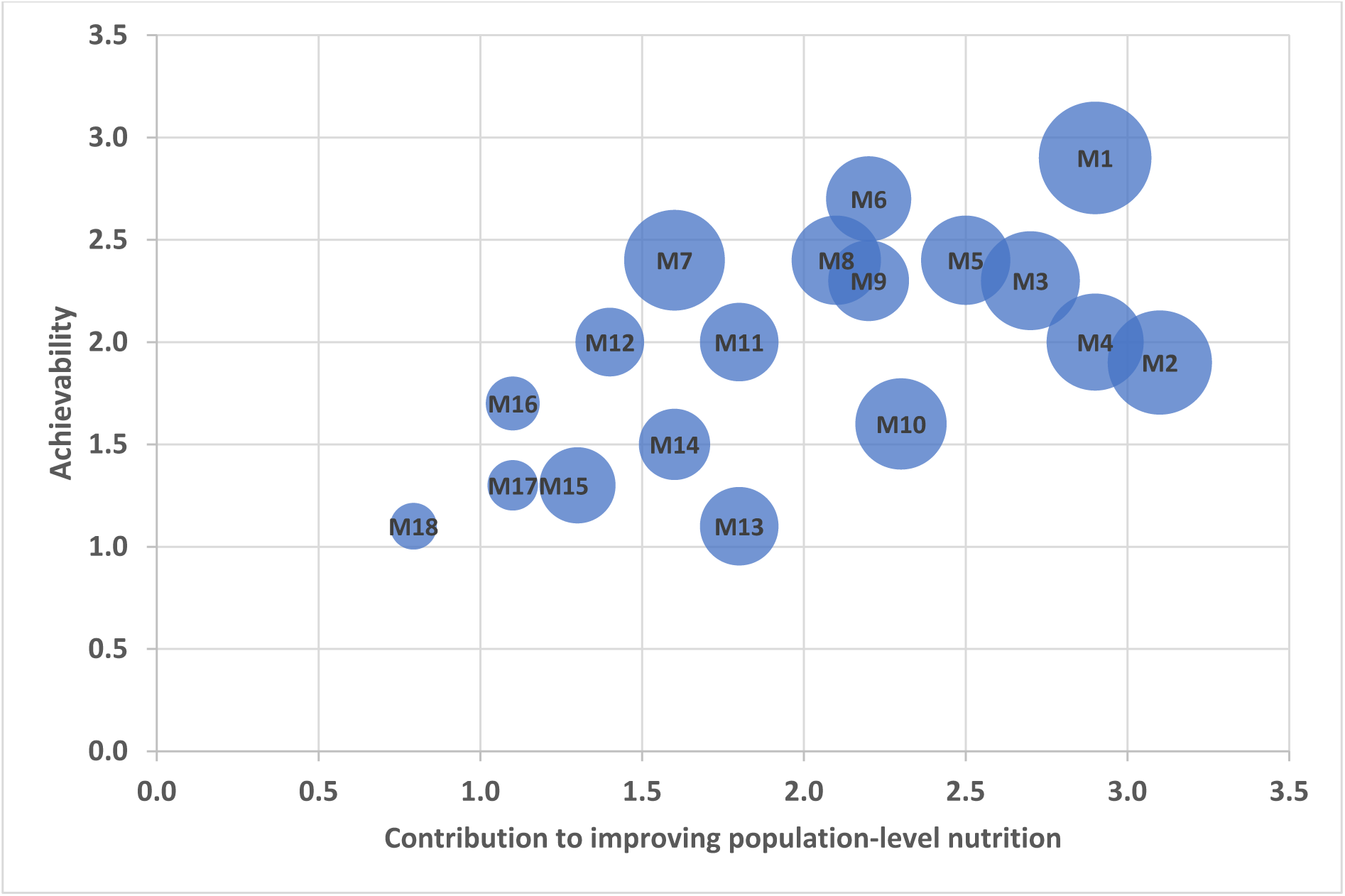
Results of the prioritization of policy actions. The codes M1-M18 are explained in table 2. The size of the dots represents the scores on criterion 3 (contribution to reducing social inequalities in nutrition and health).

For infrastructure support actions, the following recommendations were considered to have the largest potential positive impact on population-level nutrition (see table 3): evaluation of interventions and policies; monitoring of nutritional status and dietary behaviour; and knowledge transfer between policy, practice and research. The highest achievability ratings were given to the evaluation of interventions and policies, as well as local networks and initiatives.

## Discussion

The policy analysis using the Food-EPI framework reveals strengths and weaknesses of Germany’s nutrition policy landscape. The results of the international benchmarking indicate that in some policy areas Germany has strong, institutionalized structures for promoting healthy nutrition. This applies, amongst others, to the derivation of official food-based dietary guidelines and nutrition targets; the existence of publicly funded bodies with secured funding tasked with promoting healthy diets; and public commitment by political leaders. However, in other areas Germany lags significantly behind current international best practices. This is particularly the case with regards to the regulation of food advertising and marketing; the consideration of health aspects in the taxation and subsidization of foods; the promotion of healthy food supplies in retail; as well as cross-sectoral health-in-all-policies approaches.

### Policy implications

The identification and prioritization of policy recommendations indicate how policymakers could address the shortcomings of Germany’s nutrition policy framework revealed by the benchmarking. The results of our prioritization exercise are in line with existing recommendations from expert groups and professional and scientific associations, including the Scientific Advisory Board at Germany’s Federal Ministry of Food and Agriculture and the German Alliance for Non-communicable Diseases (33, 34). The mandatory, nationwide and publicly financed implementation of the existing nutrition standards for schools and kindergartens was rated the highest across the three prioritization criteria population impact, achievability and contribution to reducing inequalities. This requires government subsidies for running costs, as well as investments in canteens and dining halls. For this purpose, a federal investment programme has been proposed, which could be implemented as part of the German and EU Covid-19 recovery programme (33). The policy measures ranked the second and third highest were a health-promoting value-added tax on foods and beverages, and a tax on SSB. Although health-related food and beverages taxes – SSB taxes in particular – have gained momentum across the globe (35), so far the German government has been reluctant to consider this approach. However, it may gain traction in light of the fiscal implications of the Covid-19 pandemic, and a number of policymakers and political parties in Germany have come forward in support of the use of economic instruments in nutrition policy (36). Similarly, the policy recommendation ranked the fourth-highest – a stricter regulation of food marketing directed towards children – has recently received considerable attention from policymakers in Germany (37).

Many of the priority actions identified in our study could be complemented by action on the EU level, or would even need changes of EU regulation to be fully implemented. For example, the health-promoting VAT reform – the second-highest ranked policy recommendation in our study – would ideally involve a reduction of the VAT on healthy foods such as fruit and vegetables from the current rate in Germany of 7% to the lowest possible rate of 0%. However, EU regulation currently requires a minimum VAT rate of 5%, limiting the use of the tax system for health promotion. Indeed, in the application of the Food-EPI on the EU level, conducted as part of a separate project, the recommendation to allow Member States to lower the VAT to 0% for healthy foods was ranked third-highest (38). In general, recent analyses have found that the EU does not make full use of its potential to support healthy diets (38, 39).

The results show the potential for policy learning from international best practices. Based on the assessment of our expert panel, Germany does not reach a high level of implementation relative to international best practices for any of the 47 indicators, and very low to low levels for a majority of indicators. This shows the large potential for strengthening Germany’s policy framework for improving food environments and population-level nutrition by adopting current international best practices.

### Strengths, challenges, and limitations

The strengths of the present work include: the use of a comprehensive, rigorous, internationally harmonized methodological framework; high standards in the conduct and reporting of the research, in line with the Consolidated Criteria for Reporting Qualitative Studies (COREQ) (30); an extensive analysis of the current policy landscape in Germany and its clear and detailed presentation in the form of an evidence report; the comparison of the degree of implementation in Germany with current international best practices, i.e. with a benchmark that has already been achieved in practice in other countries; and the inclusion of the expertise of a wide circle of experts from academia, public administration, and civil society who were consulted in a multi-stage and multimodal process, as well as the high response rate.

A challenge in applying the Food-EPI framework to Germany was its federal structure with responsibilities distributed across various levels (municipalities, 16 states, the federal government, and the EU). In measuring and assessing the degree of implementation, we also considered policies, programmes and initiatives on the level of municipalities and states but less comprehensively than on the federal level. For policies and programmes on the EU level, we assessed their extent and the way they are implemented in Germany.

The results of the Food-EPI are derived from the reasoned judgement of experts based on a systematic and comprehensive compilation and analysis of relevant data. It therefore inevitably involves a certain degree of subjectivity. Alternative assessment methods, as well as a different selection of participating experts, may have produced divergent results, at least in detail. The benchmarking was carried out in July 2020 and the prioritization from November 2020 to January 2021. Some of the policy areas covered by our analysis are developing dynamically, and evaluations can therefore quickly become out of date. The Food-EPI focuses on the analysis of the influence of nutrition on human health. However, social, environmental and animal welfare aspects play an important role in food policy, too (33). Due to limited capacities and the complexity of these issues, it was not possible to consider these aspects extensively within the scope of this project, but it should be considered for future projects.

## Conclusions

The present analysis shows that the nutrition policy landscape in Germany features a number of strengths, but also substantial room for improvement. Existing policies and structures were rated as particularly low in the following areas: the regulation of food advertising, food taxation, the promotion of a healthy food supply in retail, and cross-sectoral approaches. Priority actions to address these shortcomings include a mandatory implementation of nutrition standards for schools and kindergartens; a health-promoting reform of the value added tax and a tax on sugar-sweetened beverages; as well as stricter regulation of food marketing targeting children. Efforts by a broad range of actors, including policy makers, civil society and academia, are required to support action on these priority measures for promoting healthy nutrition at the population level.

## Data Availability

All data produced in the present study are available upon reasonable request to the authors

## Acknowledgments

We would like to acknowledge the substantial and generous contributions made by the 55 external experts, listed in the supplementary appendix, who comprised the expert panel that informed the present work. We would also like to thank the Policy Evaluation Network (PEN, www.jpi-pen.eu) as well as INFORMAS (International Network for Food and Obesity/non-communicable Diseases Research, Monitoring and Action Support) for providing protocols used in this research.

## References

1. Afshin A, Sur PJ, Fay KA, et al.: Health effects of dietary risks in 195 countries, 1990-2017: a systematic analysis for the Global Burden of Disease Study 2017. The Lancet 2019; 393:1958–72.

2. NCDRisC: NCD Risk Factor Collaboration http://ncdrisc.org/.

3. IHME: Instiute for Health Metrics and Evaluation. GBD Results Tool http://ghdx.healthdata.org/gbd-results-tool (last accessed on October 10, 2019.

4. Popkin BM, Corvalan C, Grummer-Strawn LM: Dynamics of the double burden of malnutrition and the changing nutrition reality. The Lancet 2020; 395:65–74.

5. Baker P, Machado P, Santos T, et al.: Ultra-processed foods and the nutrition transition: Global, regional and national trends, food systems transformations and political economy drivers. Obesity reviews : an official journal of the International Association for the Study of Obesity 2020; 21:e13126.

6. Pingali P: Westernization of Asian diets and the transformation of food systems: Implications for research and policy. Food Policy 2007; 32:281–98.

7. Vandevijvere S, Chow CC, Hall KD, Umali E, Swinburn BA: Increased food energy supply as a major driver of the obesity epidemic: a global analysis. Bull World Health Organ 2015; 93:446–56.

8. World Health Organization: Prevalence of obesity among adults, BMI ≥ 30, age-standardized Estimates by WHO region https://apps.who.int/gho/data/view.main.REGION2480A?lang=en (last accessed on 6th of June 2021).

9. Abarca-Gómez L, Abdeen ZA, Hamid ZA, et al.: Worldwide trends in body-mass index, underweight, overweight, and obesity from 1975 to 2016: a pooled analysis of 2416 population-based measurement studies in 128·9 million children, adolescents, and adults. The Lancet 2017; 390:2627–42.

10. NCDRisk-Collaborators: Worldwide trends in diabetes since 1980: a pooled analysis of 751 population-based studies with 44 million participants. The Lancet 2016; 387:1513–30.

11. Swinburn B, Vandevijvere S, Kraak V, et al.: Monitoring and benchmarking government policies and actions to improve the healthiness of food environments: a proposed Government Healthy Food Environment Policy Index. Obesity Reviews 2013; 14:24–37.

12. Mozaffarian D, Angell SY, Lang T, Rivera JA: Role of government policy in nutrition—barriers to and opportunities for healthier eating. BMJ 2018; 361:k2426.

13. Vandevijvere S, Barquera S, Caceres G, et al.: An 11-country study to benchmark the implementation of recommended nutrition policies by national governments using the Healthy Food Environment Policy Index, 2015-2018. Obesity reviews : an official journal of the International Association for the Study of Obesity 2019.

14. Breda J, Castro LSN, Whiting S, et al.: Towards better nutrition in Europe: Evaluating progress and defining future directions. Food Policy 2020; 96:101887.

15. Hilbig A, Heuer T, Krems C, et al.: Wie isst Deutschlandã Auswertungen der Nationalen Verzehrsstudie II zum Lebensmittelverzehr. Ernährungs-Umschau : Forschung & Praxis 2009; 56:16–23.

16. Max Rubner Institut: Längsschnittstudie NEMONIT www.mri.bund.de/de/institute/ernaehrungsverhalten/forschungsprojekte/nemonit/ (last accessed on November 24, 2019.

17. OECD: OECD Health Policy Studies: The Heavy Burden of Obesity - The Economics of Prevention 2019.

18. Schienkiewitz A, Damerow S, Schaffrath Rosario A, Kurth B-M: Body-Mass-Index von Kindern und Jugendlichen: Prävalenzen und Verteilung unter Berücksichtigung von Untergewicht und extremer Adipositas. Bundesgesundheitsblatt - Gesundheitsforschung - Gesundheitsschutz 2019; 62:1225–34.

19. Hoebel J, Kuntz B, Kroll LE, et al.: Socioeconomic Inequalities in the Rise of Adult Obesity: A Time-Trend Analysis of National Examination Data from Germany, 1990–2011. Obesity Facts 2019; 12:344–56.

20. Meier T, Senftleben K, Deumelandt P, Christen O, Riedel K, Langer M: Healthcare Costs Associated with an Adequate Intake of Sugars, Salt and Saturated Fat in Germany: A Health Econometrical Analysis. PLOS ONE 2015; 10:e0135990.

21. von Philipsborn, Drees S, Geffert K, Krisam M, Nohl-Deryk P, Stratil J: Prävention und Gesundheitsförderung im Koalitionsvertrag: Eine qualitative Analyse. Das Gesundheitswesen 2018; 80:e54–e61.

22. von Philipsborn P, Drees S, Geffert K, Stratil J: Prävention von Adipositas und Diabetes mellitus: Eine Zwischenbilanz zur Mitte der Legislaturperiode. Adipositas 2019; 13:160.

23. von Philipsborn P, Effertz T, Laxy M, Schwettmann L, Stratil JM: Prävention von Adipositas und Diabetes mellitus als gesundheitspolitische Herausforderung. Adipositas 2018; 12:113–19.

24. Hawkes C, Jewell J, Allen K: A food policy package for healthy diets and the prevention of obesity and diet-related non-communicable diseases: The NOURISHING framework. Obesity reviews : an official journal of the International Association for the Study of Obesity 2013; 14:159–68.

25. Vandevijvere S, Swinburn B: Towards global benchmarking of food environments and policies to reduce obesity and diet-related non-communicable diseases: Design and methods for nation-wide surveys. BMJ open 2014; 4.

26. INFORMAS: INFORMAS Protocols http://www.informas.org/protocols/ (last accessed on 15.01.2018.

27. INFORMAS: INFORMAS Protocol: Public Sector Module. Healthy Food Environment Policy Index (Food-EPI) https://figshare.com/articles/INFORMAS_Protocol_Public_Sector_Module_-_Healthy_Food_Environment_Policy_Index_Food-EPI_/5673439 (last accessed on March 2, 2020.

28. INFORMAS: INFORMAS Countries www.fmhs.auckland.ac.nz/en/soph/global-health/projects/informas/regions.html (last accessed on 2018-08-01.

29. Lakerveld J, Woods C, Hebestreit A, et al.: Advancing the evidence base for public policies impacting on dietary behaviour, physical activity and sedentary behaviour in Europe: The Policy Evaluation Network promoting a multidisciplinary approach. Food Policy 2020: 101873.

30. Tong A, Sainsbury P, Craig J: Consolidated criteria for reporting qualitative research (COREQ): a 32-item checklist for interviews and focus groups. International Journal for Quality in Health Care 2007; 19:349–57.

31. von Philipsborn P, Geffert K, Klinger C, Hebestreit A, Stratil J, Rehfuess E: Food Environment Policy Index (Food-EPI) Evidenzbericht für Deutschland https://www.jpi-pen.eu/images/reports/Food-EPI_Germany_Evidence_Report.pdf (last accessed on 18. Mai 2021.

32. World Cancer Research Fund International: NOURISHING framework https://www.wcrf.org/int/policy/nourishing/our-policy-framework-promote-healthy-diets-reduce-obesity (last accessed on January 15, 2019.

33. Wissenschaftlicher Beirat für Agrarpolitik Ernährung und gesundheitlichen Verbraucherschutz (WBAE) beim BMEL: Politik für eine nachhaltigere Ernährung: Eine integrierte Ernährungspolitik entwickeln und faire Ernährungsumgebungen gestalten https://www.bmel.de/SharedDocs/Downloads/DE/_Ministerium/Beiraete/agrarpolitik/wbae-gutachten-nachhaltige-ernaehrung.html (last accessed on September 24, 2020.

34. Schaller K, Effertz T, Gerlach S, Grabfelder M, Müller MJ: Prävention nichtübertragbarer Krankheiten – eine gesamtgesellschaftliche Aufgabe. Grundsatzpapier der Deutschen Allianz Nichtübertragbare Krankheiten (DANK). Berlin: Deutsche Allianz Nichtübertragbare Krankheiten (DANK) 2016.

35. Popkin BM, Ng SW: Sugar-sweetened beverage taxes: Lessons to date and the future of taxation. PLoS medicine 2021; 18:e1003412.

36. von Philipsborn P, Heise TL, Lhachimi SK, Landgraf R, Hauner H: Adipositas-Prävention: Eine Steuer auf Süßgetränke ist an der Zeit. Deutsches Ärzteblatt 2017; 114:A160–4.

37. von Philipsborn P: Lebensmittel mit Kinderoptik und deren Bewerbung: Problemlage und Möglichkeiten der politischen Regulierung https://www.vzbv.de/sites/default/files/downloads/2021/02/16/vzbv_philipsborn_bericht_kindermarketing_2021-02.pdf (last accessed on February 18, 2021.

38. Djojosoeparto S, Kamphuis C, Vandevijvere S, Harrington J, Poelman M, on behalf of the JPI-HDHL Policy Evaluation Network: The Healthy Food Environment Policy Index (Food-EPI): European Union. An assessment of EU-level policies influencing food environments and priority actions to create healthy food environments in the EU https://www.jpi-pen.eu/images/reports/Food-EPI_EU_FINAL_20210305.pdf (last accessed on June 22 2021.

39. De Schutter O, Jacobs N, Clément C: A ‘Common Food Policy’ for Europe: How governance reforms can spark a shift to healthy diets and sustainable food systems. Food Policy 2020; 96:101849.

