## Supplementary Appendix for "Nutrition Policies in Germany: A Systematic Assessment with the Food Environment Policy Index (Food-EPI)"

### 1. External experts

Table s1 lists the names and affiliations of the members of the expert panel who agreed to be mentioned by name. Nine experts indicated that they preferred not to be mentioned by name. We are deeply grateful to all members for their substantial contribution of time and effort.

| Name | Affiliation (English) |
|---|---|
| **Prof. Ulrike Arens-Azevedo** | n.a. |
| **Melanie Bahlke** | Bariatric Surgery Patient Organization Germany |
| **Katrin Bindeballe** | German NCD Alliance (DANK) and German Diabetes Society |
| **Barbara Bitzer** | German NCD Alliance (DANK) and German Diabetes Society |
| **Dr. Benjamin Bodirsky** | Postdam Institute for Climate Research |
| **Brigitte Bormann** | Statutory Health Promotion Agency of the state of North Rhine Westphalia |
| **Dr. Christina Breidenassel** | German Nutrition Society |
| **Dr. Margarete Büning-Fesel** | Federal Centre for Nutrition |
| **Prof. Anette Buyken** | University of Paderborn |
| **Ricarda Corleis** | German Nutrition Society |
| **Heike Dierbach** | German NCD Alliance (DANK) and German Diabetes Society |
| **PD Dr. Tobias Effertz** | Institute for Business Law, University of Hamburg |
| **Karl Emmert-Fees** | Helmholtz Centre Munich |
| **Prof. Dr. Regina Ensenauer** | Institute for Child Nutrition, Max Rubner-Institute (federal institute for nutrition research) |
| **Sonja Fahmy** | German Nutrition Society |

| Name | Affiliation (English) |
|---|---|
| **Dr. Dietrich Garlichs** | German Diabetes Society |
| **Dr. Jasmin Geppert** | German Nutrition Society |
| **Dr. Dorle Grünewald-Funk** | n.a. |
| **Prof. Hans Hauner** | Chair of Nutritional Medicine, Technical University Munich |
| **Thomas Heise** | Leibniz Institute for Prevention Research and Epidemiology – BIPS |
| **Eva Hoffmann** | German Nutrition Society |
| **Dr. Christina Holzapfel** | Chair of Nutritional Medicine, Technical University Munich |
| **Oliver Huizinga** | Foodwatch |
| **Dr. Axel Iseke** | n.a. |
| **Dr. Irmgard Jordan** | Centre for international development and environmental research, Justus-Liebig University of Giessen |
| **Dr. Kai Kolpatzik** | Federal Association of Germany's main statutory health insurance |
| **Prof. Anja Kroke** | Technical University Fulda |
| **Prof. Katja Kröller** | Technical University Anhalt |
| **Dr. Wilfried Kunstmann** | German Medical Association |
| **Prof. Rüdiger Landgraf** | n.a. |
| **Dr. Gert Mensink** | Robert Koch Institute (Germany's national public health institute) |
| **Prof. Manfred J. Müller** | University of Kiel |
| **Anke Oepping** | National Centre for Quality in School and Kindergarten Meals |
| **Dr. Sigrid Peter** | Federal Pediatricians' Association |
| **Dr. Sandra Plachta Danielzik** | Competence Network Obesity |
| **Dr. Veronika Reisig** | Bavarian Health and Food Safety Authority |
| **Dr. Margrit Richter** | German Nutrition Society |
| **Prof. Diana Rubin** | Vivantes Health Network, Centre for Nutritional Medicine Berlin |
| **Marion Rung-Friebe** | n.a. |
| **Carolin Schäfer** | German Nutrition Society |
| **Dr. Anja Schienkiewitz** | Robert Koch Institute (Germany's national public health institute) |
| **Dr. Tatjana Schütz** | n.a. |
| **Dr. Gianni Varnaccia** | Robert Koch Institute (Germany's national public health institute) |
| **Prof. Petra Warschburger** | University of Potsdam |
| **Dr. Johannes Zeiher** | Robert Koch Institute (Germany's national public health institute) |
| **Prof. Martina de Zwaan** | German Obesity Society |

***Table s1:*** *Members of the expert panel.*

### 2. Detailed description of the policy recommendations

#### 2.1. Recommendations on policy measures

| No. | Priority action |
|---|---|
| **M1** | **Mandatory nutrition standards for schools and kindergartens**<br>Mandatory, publicly funded implementation of the nutrition standards of the German Nutrition Society (Deutsche Gesellschaft für Ernährung, DGE) in schools and kindergartens, and waiving of fees for parents. The nutrition standards include specifications for the main meals offered, other food and beverages available on the school premises, characteristics of the dining halls and the arrangement of communal meals. Furthermore, it includes the |

| No. | Priority action |
|---|---|
| | provision of sufficient financial resources (e.g., under a federal investment program), professional support, and training opportunities. |
| **M2** | **Health promoting VAT**<br>Reduced value added tax rate on healthy foods, and increased value added tax rate on unhealthy foods. |
| **M3** | **Introduction of a SSB tax**<br>Introduction of a tax on SSBs based on sugar content and use of the revenue for health promotion (e.g., improvement of catering for schools and kindergartens). |
| **M4** | **Regulation of marketing of unhealthy foods towards children**<br>Ban on advertisement of unhealthy foods directed at children, including all forms of advertising (television, internet, print and outdoor advertising, point-of-sale advertising, direct marketing, packaging, and advertising in kindergartens, schools, playgrounds and other sports and recreational facilities frequented by children) and based on an appropriate nutrient profile model (e.g., the model of the WHO European Regional Office). |
| **M5** | **Mandatory nutrition standards for public institutions (other than schools and kindergartens)**<br>Mandatory implementation of the nutrition standards of the German Nutrition Society in public institutions, such as public offices, clinics, senior citizen facilities and universities. The nutrition standards include specifications for the main meals offered, other food and beverages available on the premises, characteristics of the dining rooms and the arrangement of communal meals. Furthermore, it includes the provision of sufficient financial resources (e.g., under a federal investment program), professional support, and training opportunities. |
| **M6** | **Action plan on the promotion of drinking water**<br>Measures to promote tap water consumption, including requiring food service establishments to provide tap water free of charge or for a small service fee, offering free tap water in workplace cafeterias and canteens, and promoting tap water consumption in schools and kindergartens. |
| **M7** | **School fruit and vegetable program**<br>Expansion of the European Union's School Fruit and Vegetable Program to all German states and improved support for school authorities in its implementation. |
| **M8** | **Nutrition education in kindergartens and schools**<br>Promotion of nutrition education in kindergartens and schools by upgrading the corresponding content in the curricula of existing subjects and upgrading the teaching of home economics. |
| **M9** | **Mandatory implementation of the Nutri-Score**<br>Commitment of the German government to the EU-wide, mandatory introduction of the Nutri-Score as an interpretative, front-of-pack nutrition labelling system, accompanied by information and education of the population. |
| **M10** | **Reformulation of processed food**<br>Comprehensive and effective measures for the reformulation of processed food with the aim of a relevant reduction of nutrients of concern (sugar and other highly processed carbohydrates, salt, saturated and trans fatty acids), involving the catering sector and all relevant food groups, using binding limit values for selected food groups (including children's foods and children's dishes in restaurant chains), and with close-meshed and meaningful monitoring. |
| **M11** | **Improvement of nutritional standards for private companies**<br>Support program for the implementation of the nutritional standards of the German Nutrition Society for catering in private companies, including technical support, training offers and financial incentives (e.g., subsidies for necessary investments and certification |

| No. | Priority action |
|---|---|
| | costs). |
| M12 | **Municipal food policies**<br>Municipal food policies, including the promotion of local supply chains for fresh and low-processed foods through farmers' markets and stores, direct agricultural marketing to schools and other public institutions, establishment of school and community gardens, and establishment of decentralized lunch counters especially for the elderly, among others. |
| M13 | **Regulation of food advertising in general**<br>More effective regulation of food advertising in general, including the mandatory disclosure of the Nutri-Score in food advertising, as well as disclosure of advertising on social media. |
| M14 | **Regulations for retail outlets**<br>Rules for the placement and promotion of foods within stores, for example, rules on healthy supermarket checkouts, restrictions on special offers of non-recommended foods (esp. processed foods high in fat, sugar and/or salt). |
| M15 | **Reduction of portion and packaging sizes**<br>Measures to reduce portion and packaging sizes by regulating portion sizes in public communal catering (as part of the binding implementation of the nutritional standards of the German Nutrition Society), making them a topic in educational campaigns (e.g., the "Too good for the garbage can" campaign), initiating voluntary measures by industry, improving monitoring, and testing innovative approaches. |
| M16 | **Effective regulation of health claims**<br>Effective restriction of misleading health and nutritional claims on foods (so-called health claims), through full and consistent implementation of the Health Claims Regulation. In this context, the use of health and nutritional claims should be restricted to products with a positive overall health rating (Nutri-Score A and B), and indirect health claims (so-called "feel-good claims") that have not been regulated to date should also be covered. |
| M17 | **Mandatory nutrition labelling on restaurant menus**<br>Limited to restaurant chains with more than 20 branches and to dishes offered over a longer period of time. |
| M18 | **Quality standards for children's meals in restaurants**<br>Mandatory nutritional standards for children's meals in restaurants and snack bars, for example requiring that children's menus include water as a standard beverage and at least one vegetable or fruit component. |

***Table s2:*** *Recommendations on policy measures.*

### 2.2. Recommendations on supportive functions and structures

| No. | Priority action |
|---|---|
| S1 | **Evaluation of interventions and policies**<br>Improved evaluation of existing and planned measures for the promotion of healthy nutrition, by providing financial resources and creating appropriate structures for independent and scientifically based evaluations. |
| S2 | **Monitoring of nutritional status and dietary behaviour**<br>Improved monitoring of dietary behaviour and status through the provision of sufficient funding for regular, comprehensive, and nationally representative surveys of dietary behaviour and status (including body weight, purchasing and food preparation, food culture, and nutrition literacy), with particular attention to risk groups and reducing social inequalities. |
| S3 | **Knowledge transfer between politics, practice, and research**<br>Improved exchange of knowledge and experiences and improved cooperation between policy, |

| No. | Priority action |
|---|---|
| | practice, and science by creating appropriate structures and procedures. |
| S4 | **Nutrition education in the curricula of relevant professions**<br>Strengthen nutrition-related content in the education of relevant professional groups, including educators, teachers, physicians, medical assistants, and nurses. |
| S5 | **Monitoring of food environments**<br>Improved monitoring of food environments, including monitoring of the nutritional composition of processed foods, the extent of food advertising, food prices, and food availability in selected settings (including kindergartens, schools, universities, company cafeterias, hospitals, rehabilitation clinics, retirement homes, meals on wheels, and food banks). This includes providing sufficient financial resources for regular and close-meshed, comprehensive, and nationally representative surveys. |
| S6 | **Local networks and initiatives**<br>Strengthen initiatives and networks at the local level, including local food councils to link relevant stakeholders, and participation in the Healthy Cities Network. |
| S7 | **Action plan for healthy nutrition**<br>Develop and implement a comprehensive action plan for the promotion of healthy diets with the participation of relevant governmental and non-governmental stakeholders, considering all relevant sectors, policy, and action areas, based on a transparent, evidence-informed and reflexive process. |
| S8 | **Nutrition policy capacity building**<br>Capacity building for the development and implementation of an integrated food policy, including the institutional and budgetary upgrading of food policy in the Federal Ministry of Nutrition and Agriculture (relative to agricultural policy), the strengthening of central steering functions as well as coordination between departments and political levels, and the establishment of a consistent system of targets and indicators. |
| S9 | **Availability of data**<br>Improved accessibility to data on food environments, nutritional behaviours, and nutritional status, as well as food composition, to the public, policy makers, academia, and other stakeholders. This could be achieved, among others, with an integrated, user-friendly, freely accessible online platform as part of a national nutrition surveillance programme. |
| S10 | **Capacities for the further development of nutritional recommendations**<br>Maintaining and expanding capacities for the independent and evidence-based derivation and dissemination of nutritional recommendations by institutionally strengthening the DGE and BZfE, evaluating and evidence-based further development of existing nutritional recommendations and quality standards for community catering, taking into account nutritional and behavioural science evidence, societal objectives, and aspects of feasibility and acceptance. |

***Table s3:*** *Recommendations on supportive functions and structures.*

### 3. Adaptation of the international Food-EPI framework to Germany

For the adaptation of the international Food-EPI framework to Germany, we merged six indicators that we found to be largely overlapping in the German context into three pairs, as shown in Table s4.

| Original version (international) | Merged indicators (for Germany) |
|---|---|

<table>
<tr><td>PROV2: The government ensures that there are clear, consistent policies in other public sector settings for food service activities (canteens, food at events, fundraising, promotions, vending machines, etc.) to provide and promote healthy food choices.</td><td rowspan="2">PROV2+3: Quality and nutrition standards in other areas of the public sector: The government ensures that there are clear, consistent quality and nutrition standards for catering in other areas of the public sector (e.g., government agencies). This includes food provided in canteens and at events, as well as vending machines and other outlets in these facilities. Also included are rules for procurement.</td></tr>
<tr><td>PROV3: The government ensures that there are clear, consistent public procurement standards in public sector settings for food service activities to provide and promote healthy food choices.</td></tr>
<tr><td>GOVER3: Policies and procedures are implemented to ensure transparency in the development of food and nutrition policies.</td><td rowspan="2">GOVER3+4: Transparency and access to information: Structures and procedures are in place to ensure a high level of transparency in the development and implementation of nutrition policies. This includes public access to comprehensive nutrition information and key documents (e.g., budget documents, annual performance assessments and health indicators).</td></tr>
<tr><td>GOVER4: The government ensures public access to comprehensive nutrition information and key documents (e.g., budget documents, annual performance reviews and health indicators) for the public.</td></tr>
<tr><td>PLAT4: The government works with a system-based approach with (local and national) organisations/partners/groups to improve the healthiness of food environments at a national level.</td><td rowspan="2">HIAP1 (+PLAT4): Pursuing a systemic approach: The government takes a systemic approach to promote healthy diets and ensures that impacts on population nutrition and inequalities in nutritional status between social groups are considered in all policy areas. This includes policies such as health, agriculture, and education policies.</td></tr>
<tr><td>HIAP1: There are processes in place to ensure that population nutrition, health outcomes and reducing health inequalities or health impacts in vulnerable populations are considered and prioritised in the development of all government policies relating to food.</td></tr>
</table>

***Table s4:*** *Adaptation of the international Food-EPI framework to Germany.*

### 4. COREQ Checklist

As recommended by the *Consolidated Criteria for Reporting Qualitative Studies* (15), we provide the COREQ checklist in Table s5.

| **Domain 1: Research team and reflexivity** | |
|---|---|
| **Personal Characteristics** | |
| 1. Interviewer/facilitator: Which author/s conducted the interview or focus group? | No interviews were conducted for this study. The expert consultation including the workshop was lead by PvP and KG. |
| 2. Credentials: What were the researcher's credentials? E.g. PhD, MD | PvP: MD; MA Politics, Economics and Law; MSc Global Politics.<br>KG: MD; cand. MSc Public Health<br>CK: BSc Nutritional Sciences; MSc Public Health<br>JMS: MD; BSc Geography; cand. PhD Public |

| | |
|---|---|
| | Health and Epidemiology<br>AH: Diploma in Nutritional Sciences; PhD<br>ER: BA and MA (Oxon, Biology); PhD Public Health and Epidemiology |
| 3. Occupation: What was their occupation at the time of the study? | PvP: Research Associate and Medical Doctor<br>KG: Research Associate<br>CK: Research Associate<br>JMS: Research Associate<br>AH: Senior Scientist<br>ER: University Professor |
| 4. Gender: Was the researcher male or female? | PvP: Male<br>KG: Female<br>CK: Female<br>AH: Female<br>JMS: Male<br>ER: Female |
| 5. Experience and training: What experience or training did the researcher have? | PvP: Evidence-based public health, nutrition and health, qualitative and quantitative social science research methods.<br>KG: Evidence-based public health, nutrition and health, global health.<br>CK: Evidence-based public health, nutritional sciences, public health.<br>AH: Nutritional epidemiology, public health.<br>JMS: Social science research methods, including qualitative methods (content analysis, grounded theory); geographical research methods; public health and clinical medicine.<br>ER: Methods for evidence-based public health; evaluation of complex interventions. |
| **Relationship with participants** | |
| 6. Relationship established: Was a relationship established prior to study commencement? | Yes, to some of the participants. |
| 7. Participant knowledge of the interviewer: What did the participants know about the researcher? e.g. personal goals, reasons for doing the research | The goals of the project and the reasons for doing the research were clearly communicated to the participants (see section 6 of the online appendix). |
| 8. Interviewer characteristics: What characteristics were reported about the interviewer/facilitator? e.g. Bias, assumptions, reasons and interests in the research topic | At the beginning of the online expert workshop, all members of the research team shortly introduced themselves. |
| **Domain 2: study design** | |
| **Theoretical framework** | |
| 9. Methodological orientation and Theory: What methodological orientation was stated to underpin the study? e.g. grounded theory, discourse analysis, ethnography, phenomenology, content analysis | Policy analysis and qualitative content analysis. |

| | |
|---|---|
| **Participant selection** | |
| 10. Sampling: How were participants selected? e.g. purposive, convenience, consecutive, snowball | Purposive sampling |
| 11. Method of approach: How were participants approached? e.g. face-to-face, telephone, mail, email | By email |
| 12. Sample size: How many participants were in the study? | 55 |
| 13. Non-participation: How many people refused to participate or dropped out? Reasons? | We contacted 72 experts, of which 17 did not reply, for reasons which are unknown to us. Not all experts participated in all steps of the consultation phase, but none withdrew from their participation. |
| **Setting** | |
| 14. Setting of data collection: Where was the data collected? e.g. home, clinic, workplace | Data was collected paper-based (with questionnaires, as email text, and as comments and edits to MS Word files which were provided to the participating experts), as well as during an online workshop. |
| 15. Presence of non-participants: Was anyone else present besides the participants and researchers? | No |
| 16. Description of sample: What are the important characteristics of the sample? e.g. demographic data, date | See section 1 of the online appendix. |
| **Data collection** | |
| 17. Interview guide: Were questions, prompts, guides provided by the authors? Was it pilot tested? | n.a. |
| 18. Repeat interviews: Were repeat interviews carried out? If yes, how many? | n.a. |
| 19. Audio/visual recording: Did the research use audio or visual recording to collect the data? | The online workshop was recorded. |
| 20. Field notes: Were field notes made during and/or after the interview or focus group? | Notes were taken during the online workshop. |
| 21. Duration: What was the duration of the interviews or focus group? | The online workshop lasted for two hours. |
| 22. Data saturation: Was data saturation discussed? | In the preparation of the evidence report, we strived for data saturation. |
| 23. Transcripts returned: Were transcripts returned to participants for comment and/or correction? | A full transcript of the online workshop was produced but used internally only. A two-page summary was provided to the participants. |
| **Domain 3: analysis and findings** | |
| **Data analysis** | |
| 24. Number of data coders: How many data coders coded the data? | Two (PvP and KG). |
| 25. Description of the coding tree: Did authors provide a description of the coding tree? | We used the indicator system of the Food-EPI framework (see table 1 of the main manuscript) as a deductive coding tree. |

| | |
|---|---|
| 26. Derivation of themes: Were themes identified in advance or derived from the data? | Themes were identified in advance based on the Food-EPI framework. |
| 27. Software: What software, if applicable, was used to manage the data? | MAXQDA (Verbi GmbH, Berlin, Germany) |
| 28. Participant checking: Did participants provide feedback on the findings? | Yes (the evidence report, and various parts of the data on which the manuscript is based were provided to the participants, who were asked to provide feedback). |
| **Reporting** | |
| 29. Quotations presented: Were participant quotations presented to illustrate the themes / findings? Was each quotation identified? e.g. participant number | n.a. |
| 30. Data and findings consistent: Was there consistency between the data presented and the findings? | Yes (see results section). |
| 31. Clarity of major themes: Were major themes clearly presented in the findings? | Yes (see results section). |
| 32. Clarity of minor themes: Is there a description of diverse cases or discussion of minor themes? | Yes (see results and discussion section). |

***Table s5:*** *The COREQ checklist*

### References

1. Schaller K, Effertz T, Gerlach S, Grabfelder M, Müller MJ: Prävention nichtübertragbarer Krankheiten – eine gesamtgesellschaftliche Aufgabe. Grundsatzpapier der Deutschen Allianz Nichtübertragbare Krankheiten (DANK). Berlin: Deutsche Allianz Nichtübertragbare Krankheiten (DANK) 2016.

2. WHO Regional Office for Europe: European Food and Nutrition Action Plan 2015–2020 www.euro.who.int/en/publications/abstracts/european-food-and-nutrition-action-plan-20152020 (last accessed on November 22, 2019.

3. WHO-EURO: Vienna Declaration on Nutrition and Noncommunicable Diseases in the Context of Health 2020. WHO-EURO 2013.

4. Wissenschaftlicher Beirat für Agrarpolitik Ernährung und gesundheitlichen Verbraucherschutz (WBAE) beim BMEL: Politik für eine nachhaltigere Ernährung: Eine integrierte Ernährungspolitik entwickeln und faire Ernährungsumgebungen gestalten https://www.bmel.de/SharedDocs/Downloads/DE/_Ministerium/Beiraete/agrarpolitik/wbae-gutachten-nachhaltige-ernaehrung.html (last accessed on September 24, 2020.

5. WHO: Tackling NCDs: 'best buys' and other recommended interventions for the prevention and control of noncommunicable diseases https://apps.who.int/iris/handle/10665/259232 (last accessed on November 24, 2019.

6. vzbv und VBE: Einflussnahme von Wirtschaft in Schule: Ergebnisse einer Befragung aller Kultusministerien www.vbe.de/fileadmin/user_upload/VBE/Presse/2020-03-11_Langfassung_Sponsoring_Analyse-und-Forderungen_vzbv-und-VBE.pdf (last accessed on June 5, 2020.

7. World Health Organization: A framework for implementing the set of recommendations on the marketing of foods and non-alcoholic beverages to children

https://www.who.int/dietphysicalactivity/framework_marketing_food_to_children/en/ (last accessed on July 1, 2020.
8. Verbraucherzentrale Bundesverband (vzbv): Nationale Reduktions- und Innovationsstrategie: Mehr Tempo und strengere Vorgaben für Produkte mit Kinderoptik gefordert https://www.vzbv.de/dokument/zucker-fett-und-salzreduktion-bisher-ohne-ueberzeugende-erfolge (last accessed on July 1, 2020.
9. WHO: Follow-up to the Political Declaration of the High-level Meeting of the General Assembly on the Prevention and Control of Non-communicable DiseasesWHA 66.10 Agenda item 13.1 and 13.2).
10. Hsieh H-F, Shannon SE: Three Approaches to Qualitative Content Analysis. Qualitative Health Research 2005; 15: 1277-88.
11. Swinburn B, Vandevijvere S, Kraak V, et al.: Monitoring and benchmarking government policies and actions to improve the healthiness of food environments: a proposed Government Healthy Food Environment Policy Index. Obesity Reviews 2013; 14: 24-37.
12. INFORMAS: INFORMAS Protocol: Public Sector Module. Healthy Food Environment Policy Index (Food-EPI) https://figshare.com/articles/INFORMAS_Protocol_Public_Sector_Module_-_Healthy_Food_Environment_Policy_Index_Food-EPI_/5673439 (last accessed on March 2, 2020.
13. von Philipsborn P, Geffert K, Klinger C, Hebestreit A, Stratil J, Rehfuess E: Food Environment Policy Index (Food-EPI) Evidenzbericht für Deutschland https://www.jpi-pen.eu/images/reports/Food-EPI_Germany_Evidence_Report.pdf (last accessed on 18. Mai 2021.
14. World Cancer Research Fund International: NOURISHING framework https://www.wcrf.org/int/policy/nourishing/our-policy-framework-promote-healthy-diets-reduce-obesity (last accessed on January 15, 2019.
15. Tong A, Sainsbury P, Craig J: Consolidated criteria for reporting qualitative research (COREQ): a 32-item checklist for interviews and focus groups. International Journal for Quality in Health Care 2007; 19: 349-57.
